# Exploring the outcomes and endpoints used in gastrointestinal research in cystic fibrosis: a systematic review

**DOI:** 10.1101/2025.03.13.25323012

**Authors:** Rebecca J Calthorpe, Alexander Yule, Jemila Holaman, Sherie Smith, Helen Barr, Ryan Marsh, Charlie McLeod, Kim S Thomas, Alan R Smyth

## Abstract

**Background:** Cystic fibrosis (CF) research has increasingly focused on understanding the extra-pulmonary manifestations of CF, including on the gastrointestinal (GI) system. The effect of cystic fibrosis transmembrane conductance regulator (CFTR) modulator therapies outside the lungs is also a topic of research interest and both are key research priorities. However, significant evidence gaps persist in understanding the complex pathophysiology of CFTR dysfunction in the GI tract, and the treatment of these GI problems. Inconsistencies in outcome reporting may contribute towards these evidence gaps, and a standardised approach to outcome reporting may help to address this. This systematic rapid review aims to identify and catalogue the range of outcome measurement instruments (OMIs) and associated endpoints currently used in CF GI research.

**Methods:** This PROSPERO-registered review (CRD42021281961) was conducted following Cochrane Rapid Reviews Methods Group and COMET initiative guidance. Comprehensive searches were performed in MEDLINE, EMBASE, PubMed, Cochrane Library, and ongoing clinical trials databases, covering an 11-year period (August 2013 to November 2024). Screening and data extraction were carried out using Covidence online software.

**Results:** A total of 1,541 studies were identified, of which 193 met inclusion criteria. These studies collectively used 246 distinct OMIs, of which 172 (70%) were employed in only one study. The OMIs identified were grouped into 14 sub-domains representing key areas of GI research in CF, which were subsequently mapped to 11 of the 38 outcome domains in the taxonomy proposed by the COMET Initiative. The identified outcomes spanned a diverse range of mechanistic and patient-centred measures, reflecting the complexity of GI disease in CF.

**Conclusions:** Current research into the GI tract in CF uses a heterogeneous array of OMIs, with limited standardisation. This highlights both the complexity of CFTR dysfunction within the GI tract, requiring a wide scope of OMIs to address this, as well the variability and potential inefficiency in current outcome reporting practices. To advance our understanding of CF pathophysiology in the GI tract, a standardised approach to outcome reporting is needed. Our findings support the development of a core outcome set to promote reporting consistency and improve comparability across studies in CF GI research.

## 1. Introduction

Gastrointestinal (GI) symptoms and complications are important extra-pulmonary manifestations in cystic fibrosis (CF) yet their underlying pathophysiology is not fully understood(1). Current evidence suggests GI manifestations are multifactorial. In the upper GI tract, symptoms may arise from a combination of oesophageal dysfunction and greater negative inspiratory intrathoracic pressures(1). In the small and large intestine, a triad of GI dysmotility, microbiome dysbiosis and increased inflammation may be responsible(1). GI symptoms are common and intrusive in CF and have persisted despite the widespread introduction of cystic fibrosis transmembrane conductance regulator (CFTR) modulators(2, 3). Reducing these symptoms and understanding the extra-pulmonary effects of CFTR modulators (such as on the GI tract) have been identified as key research priorities by the CF community(4).

In clinical trials, outcome reporting can be diverse, varying from those outcomes which assess individual wellbeing, survival and adverse events; to those focused on physiological or clinical measurements; and extending to health economics and resource use(5, 6). However, in CF research, much like other areas of healthcare, outcome selection and reporting can be inconsistent and often lacks standardisation. There is an increased recognition within healthcare more broadly that previously there has been insufficient attention to the choice of outcome measures used in clinical trials. This diminishes the utility of clinical trial evidence and contributes to avoidable research waste(6–8). A systematic review of outcomes and endpoints used in pulmonary exacerbation studies in CF documented considerable heterogeneity in the outcomes and endpoints reported, ranging from patient reported outcome measures (PROMs) and clinical scores to laboratory tests(9). A systematic review looked at gaps in the evidence to support clinical decision making in CF and identified 111 evidence gaps, arising from 73 included reviews. Of these evidence gaps, 32% were attributable to inconsistent or inadequate outcome reporting(10).

Core outcome sets (COS) aim to address some of these challenges by improving outcome reporting in trials and avoiding duplication of research. While COS development has advanced in other areas of gastroenterology, as well as begun in other areas of CF research, no COS currently exists for GI-related studies in CF. The aim of this systematic rapid review is to define the current range of outcome measurement instruments (OMIs) and associated endpoints being used in research on the GI tract in people with CF (pwCF). This will provide insights into understanding the current state of GI research and lay the foundations to inform the “what” stage of COS development through determining the scope for possible outcome domains for inclusion in a COS.

## 2. Methods

This review followed Cochrane Rapid Reviews Methods Group recommendations by Garrity et al(11) and guidance in the COMET handbook used in COS development(7).

Key definitions of terms used within this manuscript are as follows:

Outcome measurement instruments (OMIs): Defined as tools used to capture and evaluate the effects of an intervention. These can include tests of biological samples, questionnaires, or other observational methods that assess the effect of outcomes such as treatment effectiveness (benefits) and side effects (risks)(7, 12). An example of an OMI is faecal calprotectin.

Endpoint: Defined as a specified parameter derived from the corresponding OMI used to assess the effect of an intervention (7, 12). For example, the difference in faecal calprotectin levels from baseline to 3 months.

Outcome sub-domains: Refers to sub-categories created in this review to give a more focused description of *what* is being measured. For example, the OMI faecal calprotectin in represented within the sub-domain “gastrointestinal inflammation”.

Outcome domain: The broader category as per the COMET taxonomy developed by Williamson/Clarke (revised) that encompasses the general area or system being measured (6). For example, “gastrointestinal outcomes” represents a distinct outcome domain to which gastrointestinal inflammation can be mapped.

### 2.3 Search Strategy and Inclusion Criteria

Members of the research team for this systematic rapid review represented key stakeholders in CF research including health professionals and researchers in CF, researchers with experience of core outcome set methodology and a Cochrane CF systematic reviewer. The review protocol was agreed a priori by the research team and registered on PROSPERO database (CRD42021281961) (13).

Inclusion criteria were studies of any study design that reported OMIs and associated endpoints relating to GI symptoms, complications, or pathophysiology in pwCF. Full text articles and conference abstracts were considered for inclusion where meaningful data could be extracted to capture new or emerging outcomes that had not yet reached full publication. The publication language was limited to English. The population for inclusion was a person of any age with a confirmed diagnosis of CF in accordance with the Cystic Fibrosis Foundation consensus guidelines (17).

Phase I/II trials, pharmacokinetic studies and case series containing less than 10 patients were excluded as it was felt they would be less likely to contain clinically meaningful outcome measures. Systematic reviews were also excluded as the aim was to identify and catalogue all OMIs currently used in CF research, and not those deemed important by previous systematic reviewers. Outside the scope of this review were CF complications which may result from pancreatic dysfunction such as pancreatitis or those which do not impact on the lumen of the GI tract such as CF-related diabetes. Studies relating to hepatology were also excluded.

Searches were conducted in MEDLINE, EMBASE, PubMed and the Cochrane databases over an 11-year period (August 2013 to November 2024). These date restrictions were used to include studies pre and post the widespread introduction of CFTR modulators in 2019 (14). The full search strategy is detailed in supplementary material 1 and was developed by AY with confirmation by a Cochrane CF systematic reviewer (SS). Additionally, ClinicalTrials.gov, ISRCTN and EU clinical trials registries were searched to identify any ongoing clinical trials.

### 2.4 Study Screening and Data Extraction

Screening and data extraction were conducted using the online systematic review platform, Covidence. For both title/abstract and full-text reviews, 10% of studies were screened by two reviewers (AY and JH or RC and JH) using standardised pre-designed forms to establish consensus on studies for inclusion with RC and AY screening the remaining studies and JH verifying the excluded studies and resolving any conflicts. Relevant data on the included studies were extracted by RC or AY using a pilot data extraction form with 10% checked by JH, and included details such as author, journal, study title, OMIs, and associated endpoints. Risk of bias and evidence grading were not assessed because the focus of the review was to describe what outcomes and endpoints were used, rather than to evaluate the quality of studies or characteristics and measurement properties of the OMIs. The latter typically occurs once the initial core outcome set has been established.

Data were organised using Stata and OMIs were reviewed. Where different names were used, at different times, for the same instrument (e.g. JensAbdomen-CF and CFAbd-Score) these were combined into one OMI. Similar OMIs which used different metrics to report the outcome such as body mass index (BMI) percentile vs BMI z-score were considered as separate OMIs. Due to the heterogeneity of data and large number of OMIs captured, descriptive analysis on the number and percentage of times an OMI and associated endpoint was used were provided for the top 10 most commonly used OMIs.

OMIs with similar underlying functions (such as assessment of gastrointestinal inflammation) were then grouped into sub-domains and definitions describing each of the subdomains were agreed upon by the reviewers, RC and AY. The wider research team resolved difficulties and made final decisions as required. The number of times an OMI was mapped to a sub-domain, as well as the number and percentage of articles that used a particular sub-domain, were recorded. These sub-domains were then mapped to the 38-domain taxonomy by the COMET Initiative developed by Williamson/Clarke (revised) (6).

## 3. Results

Searches identified a total of 1541 articles and protocols, with 193 included in the review (supplementary material 2). Figure 1 summarises the number of articles included at each stage and reasons for exclusion.

**Figure 1.**
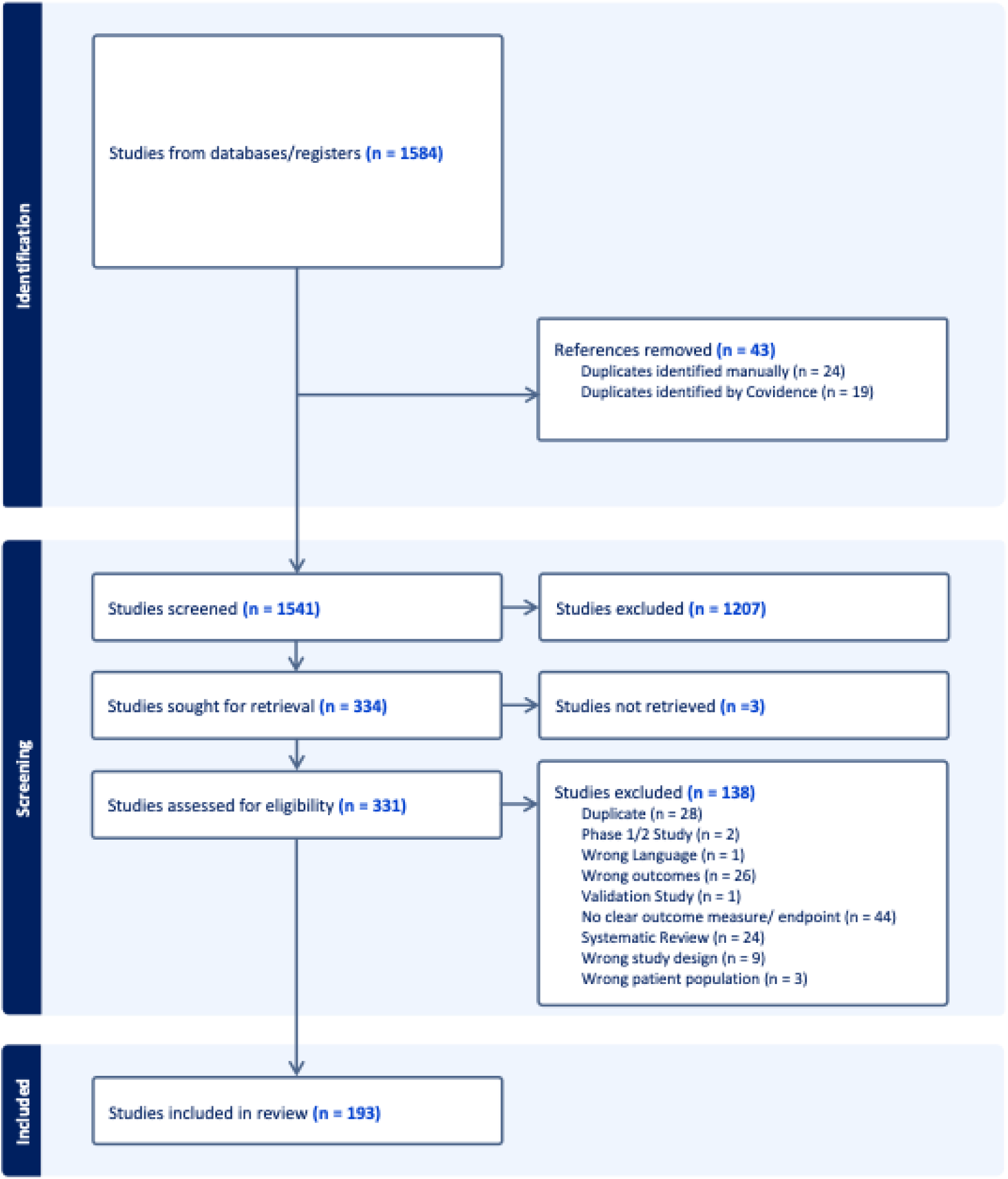
PRISMA flow chart of included and excluded studies

### Outcome Measurement Instruments

In total, 246 different OMIs were identified. A full list of OMIs and their associated sub-domains can be found in supplementary material 3. Of the 246 different OMIs identified, 172 (70%) were reported in only a single article.

The 10 most commonly reported OMIs used in the included studies were faecal calprotectin, BMI, BMI z-score, weight z-score, faecal elastase, height z-score, CFAbd-Score, Cystic Fibrosis Questionnaire-Revised (CFQ-R), Patient Assessment of Constipation - Symptoms (PAC-SYM) and weight. (Table 1).

**Table 1.** The top 10 most used OMIs in GI research in CF and details of the number of times and percentage of articles they were used in. The type of OMI and how they map to a particular sub-domain are also described.

| Rank | Outcome measurement instrument | Number times used (n) and as percentage (%) of the number articles | Type of outcome measurement instrument | Outcome sub-domain |
| --- | --- | --- | --- | --- |
| 1 | Faecal calprotectin | n=51, 26% of articles | Biomarker | Gastrointestinal inflammation |
| 2 | BMI | n=37, 19% of articles | Anthropometric measurement | Growth and body composition |
| 3 | BMI z-score | n=35, 18% of articles | Anthropometric measurement | Growth and body composition |
| 4 | Weight z-score | n=28, 15% of articles | Anthropometric measurement | Growth and body composition |
| 5 | Faecal elastase | n=25, 13% of articles | Biomarker | Exocrine pancreatic function |
| 6 | Height z-score | n=22 11% of articles | Anthropometric measurement | Growth and body composition |
| 7 | CFAbd-Score | n=20 10% of articles | Patient reported outcome measure | Gastrointestinal symptoms AND Health related quality of life |
| 8 | Cystic Fibrosis Questionnaire-Revised (CFQ-R) | n=16, 8% of articles | Patient reported outcome measure | Health related quality of life |
| 9 | Patient Assessment of Constipation – Symptoms (PAC-SYM) | n=14, 7% of articles | Patient reported outcome measure | Gastrointestinal symptoms |
| 10 | Weight | n=11 6% of articles | Anthropometric measurement | Growth and body composition |

Faecal calprotectin was the most frequently used OMI, used for assessing GI inflammation (used 51 times in 193 articles, 26%). Its use as an endpoint included to measure changes in faecal calprotectin levels between baseline and post-treatment, often in response to interventions such as CFTR modulators, including both ivacaftor and elexacaftor / tezacaftor / ivacaftor (ETI). Levels of faecal calprotectin were also compared between groups, such as intervention and control groups or pwCF and healthy controls. Additionally, levels of faecal calprotectin were correlated with patient-reported outcomes such as the CFQ-R or CFAbd-Score, clinical biomarkers, lung function and growth metrics such as height and weight. The other OMI biomarker within the top 10 was faecal elastase, used as a measure of pancreatic exocrine insufficiency.

Multiple growth-related parameters were reported within the top 10 OMIs. For weight, metrics included absolute weight and weight z-scores. Similarly, BMI was assessed using standard BMI and BMI z-scores, while height was measured as height z-scores. Across all included articles, a total of six metrics were identified for weight, five for BMI, and four for height (supplementary material 3). Three PROMs were among the top 10 OMIs identified. Two were CF-specific: the CFAbd-Score, a 28-item PROM assessing GI symptoms and quality of life, and the CFQ-R, a global health-related quality of life PROM. Additionally, a further non-CF specific PROM, PAC-SYM was also commonly used to assess lower GI symptoms.

Endpoints used across the studies included evaluating changes in OMI values pre-and post-intervention (e.g. CFTR modulators), comparisons between study groups (e.g., pwCF versus healthy controls or intervention versus control groups), and correlations between OMIs and other clinical outcomes. For PROMs, endpoints frequently involved changes in total or domain scores over time, group comparisons, and correlations with other outcomes such as GI biomarkers (e.g., faecal calprotectin, IL-1β, or M2-pyruvate kinase), imaging results (e.g., ultrasound or MRI), and timing of pancreatic enzyme replacement therapy (PERT). Additionally, PROM scores were correlated with other PROMs, such as the GI symptom tracker, Patient Health Questionnaire-9 (PHQ-9), and Generalised Anxiety Disorder-7 (GAD-7). Overall, 23 recognised PROMs were utilised across the studies to evaluate upper and lower GI symptoms, quality of life, dietary intake, and fatigue.

### Outcome Sub-Domains and mapping to the COMET taxonomy

These 246 OMIs were mapped to 14 GI outcome sub-domains. Definitions for each sub-domain and frequency of use are described in Table 2.

**Table 2.** Gastrointestinal outcome sub-domains and their definitions. Of note, a single article may be mapped to multiple sub-domains depending upon what OMIs are used.

| GI Outcome Sub-domain | Definition | Mechanistic vs Patient-Centred/Clinical | Number times used, number and percentage of included articles OMI used in |
| --- | --- | --- | --- |
| Growth and body composition | Outcomes related to growth and development in children or physical characteristics (e.g., height, weight) measured at any age. | Patient-Centred/Clinical | 199 times<br>86 articles (45% articles) |
| Gastrointestinal microbiota | Outcomes related to the presence, composition, or function of microorganisms within the lumen of the gastrointestinal tract. | Mechanistic | 92 times<br>43 articles (22% articles) |
| Gastrointestinal symptoms | Outcomes based on patient-reported symptoms related to GI function, such as bloating, flatulence, steatorrhea, borborygmi, and pain. | Patient-Centred/Clinical | 85 times<br>60 articles (31% articles) |
| Gastrointestinal inflammation | Outcomes related to inflammation and associated damage in the gastrointestinal tract. | Mechanistic | 69 times<br>52 articles (27% articles) |
| Health related quality of life | Outcomes addressing how GI function and symptoms affect daily activities, social interactions, and overall quality of life, including schooling and work. | Patient-Centred/Clinical | 53 times<br>48 articles (25% articles) |
| Exocrine pancreatic function | Outcomes related to exocrine function of the pancreas such as the secretion of pancreatic enzymes and bicarbonate required for digestion. | Mechanistic | 46 times<br>35 articles (18% articles) |
| Gastrointestinal motility | Outcomes assessing the movement and transit time of contents through the gastrointestinal tract. | Mechanistic | 46 times<br>22 articles (11% articles) |
| Gastrointestinal absorption, digestion, and emulsification | Outcomes focusing on the breakdown, processing, and absorption of intraluminal gastrointestinal contents. | Mechanistic | 34 times<br>23 articles (12% articles) |
| Oesophageal and GI intraluminal environment | Outcomes assessing conditions within the GI tract (upper and lower), such as intraluminal pH, excluding microbiota. | Mechanistic | 24 times<br>10 articles (5% articles) |
| Gastrointestinal structure | Outcomes evaluating the anatomy or structural abnormalities of GI tract | Mechanistic | 21 times<br>16 articles (8% articles) |
| Complication of cystic fibrosis | Outcomes addressing GI-related complications of CF, such as meconium ileus or distal intestinal obstruction syndrome (DIOS). | Patient-Centred/Clinical | 17 times<br>11 articles (6% articles) |
| Pancreatic structure | Outcomes assessing the anatomy or structural abnormalities of the pancreas. | Mechanistic | 16 times<br>8 articles (4% articles) |
| CF medication use | Outcomes related to the use and rates of medications specifically prescribed for CF management. | Patient-Centred/Clinical | 6 times,<br>5 articles (3% articles) |
| Diet and dietary change | Outcomes examining dietary interventions or modifications (e.g., high-fat, high-calorie diets) associated with CF management. | Patient-Centred/Clinical | 3 times<br>3 articles (2% articles) |

The most frequently assessed outcome sub-domain was growth and body composition (used 199 times across 86 articles, representing 45% of included articles), followed by gastrointestinal microbiota (92 instances across 43 articles, 22% of articles) and gastrointestinal symptoms (85 instances across 60 articles, 31% of articles) (Table 2). Sub-domains were classified as either patient-centred/clinical or mechanistic. Six patient-centred/clinical sub-domains were identified, encompassing outcomes that reflect participants’ experiences, such as symptoms, complications, quality of life and growth and body composition. In contrast, eight mechanistic sub-domains were identified, focusing on structural or physiological processes not directly perceived by participants. A description of how each OMI is mapped to a particular sub-domain can be found in supplementary material 3.

The GI outcome sub-domains were mapped to 11 of the 38 COMET taxonomy domains(6). Figure 2 summarises how the GI outcome sub-domains map to the taxonomy. Some sub-domains were able to be mapped to more than one taxonomy domain. The most commonly used COMET taxonomy domain was gastrointestinal outcomes, followed by metabolic and nutritional outcomes.

**Figure 2.**
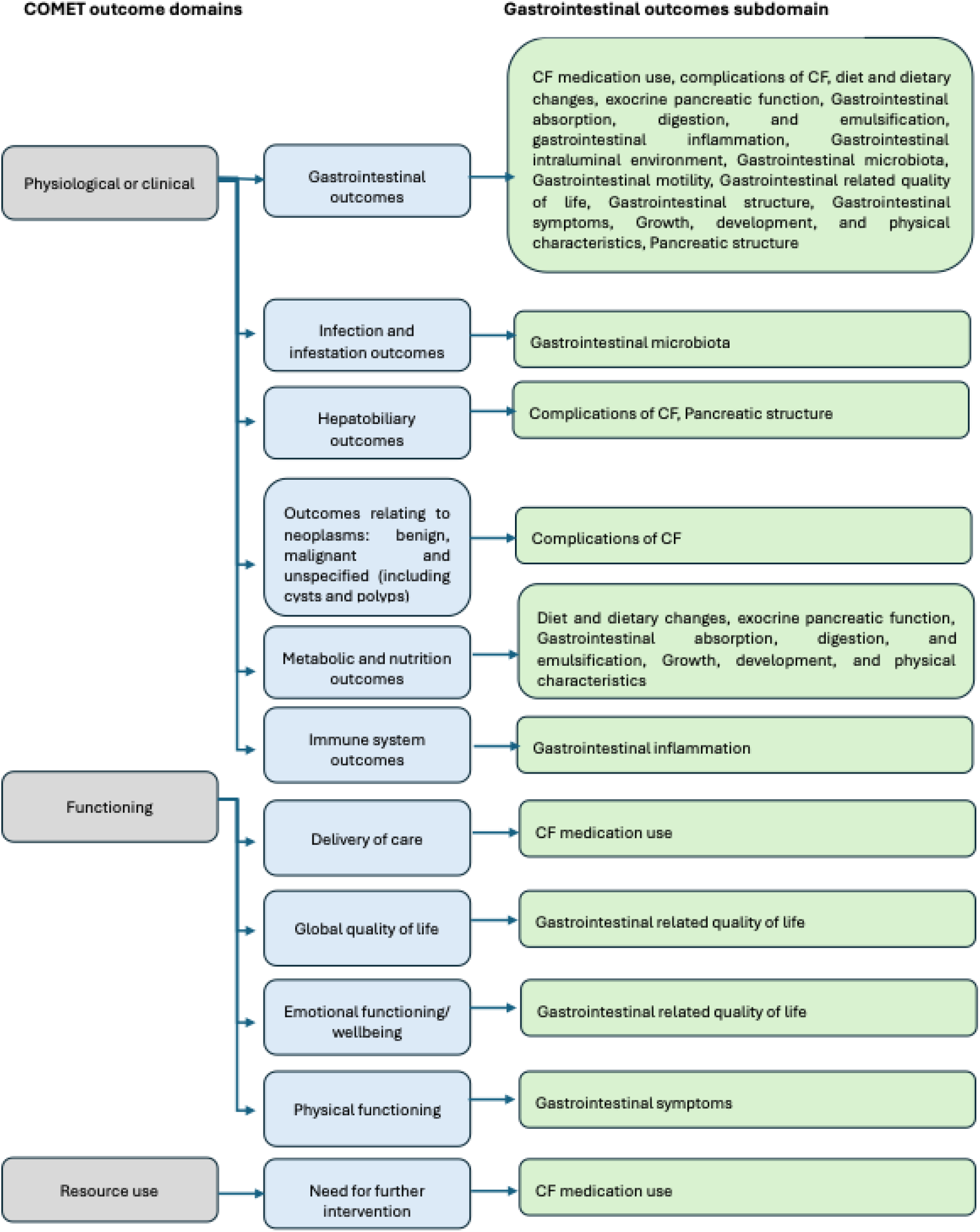
Demonstrates how each of the proposed sub-domains map to the COMET taxonomy by Williamson/Clarke (revised).

## 4. Discussion

This systematic rapid review explored the range of outcome measurements and associated endpoints used in GI research in CF. The findings revealed a heterogeneous use of OMIs within current research. Among the 193 included articles, 246 distinct OMIs were used, with 172 (70%) being reported once, highlighting the significant variability in OMI selection and use across studies. The reasons for this are likely multifaceted and in part related to the complexity of the GI tract physiology, and the GI tract’s intrinsic relationship with CFTR protein function and therefore pathophysiology in CF. Thus, a wide scope of OMIs is likely needed in order to address this. Furthermore, some OMIs, for example imaging metrics, are exploratory and can be important for identifying new ways of studying the GI tract.

However, inconsistencies in the use and reporting of common OMIs such as height, weight, and BMI, also highlights this heterogeneity likely relates to how outcomes are reported, and therefore the challenges for comparing and synthesising findings using these OMIs across different studies. The issue in the variability in OMIs used reflects the finding from other CF outcome reporting research, where a wide range of outcome reporting was also found in studies of pulmonary exacerbations in CF (9). This variability is also problematic within wider healthcare research and contributes to research inefficiency and waste(6, 8, 15).

To address these challenges, establishing a COS for GI research in CF is essential. A COS is defined as “an agreed standardised set of outcomes that should be measured and reported, as a minimum, in all clinical trials in specific areas of health or health care”(16). While two COS initiatives for CF are currently under development - one universal and another focused on pulmonary exacerbations (9, 17) no COS for GI outcomes in CF are currently registered with the COMET initiative. Notably, eight of the top 10 OMIs identified in this review were patient focused, focusing on areas such as growth and body composition, GI symptomatology and quality of life. The remaining two were biomarkers; faecal calprotectin (a marker of GI inflammation) and faecal elastase (a marker of exocrine pancreatic function). These findings suggest that current research prioritises both understanding the causes of GI dysfunction in CF as well as addressing GI symptoms and associated burden. To be comprehensive, a COS in GI research in CF will likely need to encompass both aspects.

The success of COS initiatives in other healthcare fields, such as the OMERACT collaboration in rheumatology(18), demonstrates the value of outcome standardisation. To date, there are 589 COSs reported from 480 studies (published up to 2021) developed across various disciplines(5), with 74 COSs related to gastroenterology listed in the COMET database(17). This indicates that creating a GI-specific COS for CF is both achievable and necessary to improve consistency in outcome reporting.

A strength of this review is its comprehensive scope, encompassing a wide range of studies to provide a detailed analysis of OMIs currently used in GI CF research and how they can be mapped to sub-domains and the COMET taxonomy, providing the foundational groundwork for COS development in CF GI research. Future work will need to establish which of these should be included within a COS through wider CF community involvement and reach consensus via eDelphi methods(7). Once a COS has been agreed, we would recommend following PRISMA-COSMIN guidance for the assessment of the measurement properties, certainty of the evidence of each OMI assessed, as well as risk of bias for the studies when determining how these outcomes should be measured(19, 20).

Limitations include a lack of studies addressing the economic and societal impact of GI disease in CF, despite its well documented burden, such as missed work or school days. Additionally, reporting on the number of OMIs in articles, rather than unique studies, may have led to an overestimation of OMI usage when the same study was included as both an abstract and a full-text manuscript. However, as the intended outcome measures stated in a study registration or protocol may differ from the reporting in the final manuscript of studies, reporting per article was chosen.

In conclusion, understanding CFTR function and the consequences of dysfunction in the GI tract is complex, and this review demonstrates the significant variability in outcome measurements used with GI research at present. In order to advance our understanding of CF pathophysiology in the GI tract a consistent and standardised approach to outcome reporting is needed. Creation of a COS using sub-domains described in this review as part of the development process, would be positive step towards this goal.

## Abbreviations

BMI: Body mass index
CF: Cystic Fibrosis
COS: Core outcome set
COMET Initiative: Core Outcome Measures in Effectiveness Trials Initiative
CFTR: Cystic fibrosis transmembrane regulator
DIOS: Distal intestinal obstruction syndrome
ETI: Elexacaftor / tezacaftor / ivacaftor
GI: Gastrointestinal
OMIs: Outcome measurement instruments
OMERACT: Outcome Measures in Rheumatology
PROM: Patient reported outcome measure
pwCF: People with cystic fibrosis

## Availability of data and materials

The dataset supporting the conclusions of this article is available upon reasonable request to the study chief investigator Prof Alan Smyth.

