## Supplementary material 1 for "Exploring the outcomes and endpoints used in gastrointestinal research in cystic fibrosis: a systematic review"

### Database Searches

The search strategy was intentionally broad and was designed to capture a wide range of outcome measures. Using Boolean operators, we structured the search as (cystic fibrosis AND gastrointestinal) AND (patient reported outcome measure OR outcome measures) to ensure comprehensive coverage of outcome measures in CF. Specific outcomes listed used the OR operator. This was to ensure known key outcomes were included within the search, without excluding unlisted outcomes.

Following the search update for the period August 2023 – November 2024 an additional search of (Cystic Fibrosis [AND] Gastrointestinal) was completed. A preliminary screening of these were completed to ensure that no key references had been omitted from the search when specific outcome measurement instruments were also applied within the search strategy. This did not reveal any new articles for inclusion not already highlighted using the search strategies listed below.

### EMBASE SEARCH

Using the “keyword” and “abstract’ search functions, proposed search:

(Cystic Fibrosis [AND] Gastrointestinal) [AND] patient reported outcome measures, [OR] outcome measures;

[OR]

(Cystic Fibrosis [AND] Gastrointestinal) [AND] distal intestinal obstruction syndrome, [OR] DIOS, [OR] meconium ileus, [OR] pancreatic exocrine insufficiency ,[OR] bowel inflammation;

[OR]

(Cystic Fibrosis [AND] Gastrointestinal) [AND] CFAbd-score, [OR] CFQoL, [OR] JenAbdomen-CF, [OR] PEI-Q, [OR] PAC-SYM, [OR] PAC-QOL, [OR] PAGI-SYM, [OR] PAGI-QOL, [OR] ROME IV, [OR] Bristol Stool Chart, [OR] GSRS, [OR] IBS-SS, [OR] abdominal symptom score;

[OR]

(Cystic Fibrosis [AND] Gastrointestinal) BMI, [OR] height, [OR] weight;

[OR]

(Cystic Fibrosis [AND] Gastrointestinal) [AND] orocaecal transit time, [OR] small bowel water content, [OR] total colonic volume, [OR] Whole bowel transit time, [OR] gastric half emptying time, [OR] intestinal pH, [OR] lactose ureide breath test, [OR] cholecystokinin,

[OR] pancreatic enzyme measurement, [OR] coefficient of fat absorption, [OR] 4-cholesten-3-one, [OR] serum immunoreactive trypsinogen, [OR] fat malabsorption blood test, [OR] faecal elastase-1, [OR] faecal calprotectin, [OR] magnetic resonance imaging, [OR] capsule endoscopy, [OR] ultrasound scan, [OR] computed tomography, [OR] X-ray.

Original search save (Aug 2013 – Aug 2023):
https://ovidsp.ovid.com/ovidweb.cgi?T=JS&NEWS=N&PAGE=main&SHAREDSEARCHID=1csyqKIaET80UxAECXDs34c1JNgKPbCA3XZoQxqPhFOPgIFOgPP60hk2rxESW9fEf

Updated search save (August 2023 – November 2024):

https://ovidsp.dc1.ovid.com/ovid-new-b/ovidweb.cgi

### Medline SEARCH

Using the “keyword” and “abstract’ search functions, proposed search:

(Cystic Fibrosis [AND] Gastrointestinal) [AND] patient reported outcome measures, [OR] outcome measures;

[OR]

(Cystic Fibrosis [AND] Gastrointestinal) [AND] distal intestinal obstruction syndrome, [OR] DIOS, [OR] meconium ileus, [OR] pancreatic exocrine insufficiency or exocrine pancreatic insufficiency/ ,[OR] bowel inflammation;

[OR]

(Cystic Fibrosis [AND] Gastrointestinal) [AND] CFAbd-score (patient reported outcome meaures/ or “surveys and questionnaires”, [OR] CFQoL, [OR] JenAbdomen-CF, [OR] PEI-Q, [OR] PAC-SYM, [OR] PAC-QOL, [OR] PAGI-SYM, [OR] PAGI-QOL, [OR] ROME IV, [OR] Bristol Stool Chart, [OR] GSRS, [OR] IBS-SS, [OR] abdominal symptom score;

[OR]

(Cystic Fibrosis [AND] Gastrointestinal)  BMI (or body mass index), [OR] height (or body height), [OR] weight (weights and measures);

[OR]

(Cystic Fibrosis [AND] Gastrointestinal)  [AND] orocaecal transit time (or gastrointestinal motility or gastrointestinal transit), [OR] small bowel water content, [OR] total colonic volume, [OR] Whole bowel transit time, [OR] gastric half emptying time (or gastric emptying), [OR] intestinal pH, [OR] lactose ureide breath test (or breath test), [OR] cholecystokinin or cholecystokinin/, [OR] pancreatic enzyme measurement, [OR] coefficient of fat absorption, [OR] 4-cholesten-3-one, [OR] serum immunoreactive trypsinogen, [OR] fat malabsorption blood test, [OR] faecal elastase, [OR] faecal calprotectin, [OR] magnetic resonance imaging or MRI , [OR] capsule endoscopy or capsule endoscopy/, [OR] ultrasound scan (or ultrasonography), [OR] computed tomography, [OR] X-ray or x-rays, [OR] microbiota, [OR] terminal ileum motility

Search save:
https://ovidsp.ovid.com/ovidweb.cgi?T=JS&NEWS=N&PAGE=main&SHAREDSEARCHID=13YEqQGXTFHWYvk8CntDovhJQBwK9euGopkX1u2MOLd53wmjsiPZXSugPWES2m2zu

### PubMed Search

Using the title/abstract search function:

("cystic fibrosis"[Title/Abstract] AND "gastrointestinal"[Title/Abstract] AND ("outcome measures"[Title/Abstract] OR "patient reported outcome measures"[Title/Abstract])) OR ("cystic fibrosis"[Title/Abstract] AND "gastrointestinal"[Title/Abstract] AND ("distal intestinal obstruction syndrome"[Title/Abstract] OR "meconium ileus"[Title/Abstract] OR "cystic fibrosis related liver disease"[Title/Abstract] OR "pancreatic exocrine insufficiency"[Title/Abstract] OR "bowel inflammation"[Title/Abstract])) OR ("cystic fibrosis"[Title/Abstract] AND "gastrointestinal"[Title/Abstract] AND ("height"[Title/Abstract] OR "weight"[Title/Abstract] OR "BMI"[Title/Abstract])) OR ("cystic fibrosis"[Title/Abstract] AND "gastrointestinal"[Title/Abstract] AND ("orocecal transit time"[Title/Abstract] OR "small bowel water content"[Title/Abstract] OR "whole bowel transit time"[Title/Abstract] OR "gastric half emptying time"[Title/Abstract] OR "total colonic volume"[Title/Abstract] OR "intestinal ph"[Title/Abstract] OR "lactose ureide breath test"[Title/Abstract] OR "cholecystokinin"[Title/Abstract] OR "pancreatic enzyme measurement"[Title/Abstract] OR "coefficient of fat absorption"[Title/Abstract] OR "4-cholesten-3-one"[Title/Abstract] OR "serum immunoreactive trypsinogen"[Title/Abstract] OR ("fat"[All Fields] AND "malabsorption blood test"[Title/Abstract]) OR "fecal elastase"[Title/Abstract] OR "fecal calprotectin"[Title/Abstract] OR "MRI"[Title/Abstract] OR "capsule endoscopy"[Title/Abstract] OR "ultrasound scan"[Title/Abstract] OR "CT"[Title/Abstract] OR "x-ray"[Title/Abstract] OR “microbiota”[Title/abstract] OR “fasting terminal ileum motility”[Title/Abstract])) OR (("CFAbd-score"[Title/Abstract] OR "CFQoL"[Title/Abstract] OR "JenAbdomen-CF"[Title/Abstract] OR "PAC-SYM"[Title/Abstract] OR “PAC-QOL”[Title/Abstract] OR “PAGI-SYM”[Title/Abstract] OR “PAGI-QOL”[Title/Abstract] OR “PEI-Q”[Title/Abstract] OR “ROME IV”[Title/Abstract] OR “Bristol Stool Chart”[Title/Abstract] OR “GSRS”[Title/Abstract] OR “IBS-SS”[Title/Abstract] OR "abdominal symptom score"[Title/Abstract]) AND ("cystic fibrosis"[Title/Abstract] AND "gastrointestinal"[Title/Abstract]))

### Cochrane Database Search

ID Search

#1 (cystic fibrosis):ti,ab,kw AND (gastrointestinal):ti,ab,kw AND (patient reported outcome measures):ti,ab,kw (Word variations have been searched)

#2 (cystic fibrosis):ti,ab,kw AND (gastrointestinal):ti,ab,kw AND (outcome measures):ti,ab,kw (Word variations have been searched)

#3 (cystic fibrosis):ti,ab,kw AND (gastrointestinal):ti,ab,kw AND (distal intestinal obstruction syndrome):ti,ab,kw (Word variations have been searched)

#4 (cystic fibrosis):ti,ab,kw AND (gastrointestinal):ti,ab,kw AND (meconium ileus):ti,ab,kw (Word variations have been searched)

#5 (cystic fibrosis):ti,ab,kw AND (gastrointestinal):ti,ab,kw AND (pancreatic exocrine insufficiency):ti,ab,kw (Word variations have been searched)

#6 (cystic fibrosis):ti,ab,kw AND (gastrointestinal):ti,ab,kw AND (bowel inflammation):ti,ab,kw (Word variations have been searched)

#7 (cystic fibrosis):ti,ab,kw AND (gastrointestinal):ti,ab,kw AND (height):ti,ab,kw (Word variations have been searched)

#8 (cystic fibrosis):ti,ab,kw AND (gastrointestinal):ti,ab,kw AND (weight):ti,ab,kw (Word variations have been searched)

#9 (cystic fibrosis):ti,ab,kw AND (gastrointestinal):ti,ab,kw AND ("body-mass index"):ti,ab,kw (Word variations have been searched)

#10 (cystic fibrosis):ti,ab,kw AND (gastrointestinal):ti,ab,kw AND ("orocaecal transit time"):ti,ab,kw (Word variations have been searched)

#11 (cystic fibrosis):ti,ab,kw AND (gastrointestinal):ti,ab,kw AND (small bowel water content):ti,ab,kw (Word variations have been searched)

#12 (cystic fibrosis):ti,ab,kw AND (gastrointestinal):ti,ab,kw AND (whole bowel transit time):ti,ab,kw (Word variations have been searched)

#13 (cystic fibrosis):ti,ab,kw AND (gastrointestinal):ti,ab,kw AND ("gastric half emptying time"):ti,ab,kw (Word variations have been searched)

#14 (cystic fibrosis):ti,ab,kw AND (gastrointestinal):ti,ab,kw AND (total colonic volume):ti,ab,kw (Word variations have been searched)

#15 (cystic fibrosis):ti,ab,kw AND (gastrointestinal):ti,ab,kw AND (intestinal pH):ti,ab,kw (Word variations have been searched)

#16 (cystic fibrosis):ti,ab,kw AND (gastrointestinal):ti,ab,kw AND (lactose ureide breath test):ti,ab,kw (Word variations have been searched)

#17 (cystic fibrosis):ti,ab,kw AND (gastrointestinal):ti,ab,kw AND (cholecystokinin):ti,ab,kw (Word variations have been searched)

#18 (cystic fibrosis):ti,ab,kw AND (gastrointestinal):ti,ab,kw AND (pancreatic enzyme measurement):ti,ab,kw (Word variations have been searched)

#19 (cystic fibrosis):ti,ab,kw AND (gastrointestinal):ti,ab,kw AND (coefficient of fat absorption):ti,ab,kw (Word variations have been searched)

#20 (cystic fibrosis):ti,ab,kw AND (gastrointestinal):ti,ab,kw AND (serum immunoreactive trypsinogen):ti,ab,kw (Word variations have been searched)

#21 (cystic fibrosis):ti,ab,kw AND (gastrointestinal):ti,ab,kw AND (malabsorption blood test):ti,ab,kw (Word variations have been searched)

#22 (cystic fibrosis):ti,ab,kw AND (gastrointestinal):ti,ab,kw AND ("faecal elastase-1"):ti,ab,kw (Word variations have been searched)

#23 (cystic fibrosis):ti,ab,kw AND (gastrointestinal):ti,ab,kw AND (faecal calprotectin):ti,ab,kw (Word variations have been searched)

#24 (cystic fibrosis):ti,ab,kw AND (gastrointestinal):ti,ab,kw AND ("magnetic resonance imaging"):ti,ab,kw (Word variations have been searched)

#25 (cystic fibrosis):ti,ab,kw AND (gastrointestinal):ti,ab,kw AND (capsule endoscopy):ti,ab,kw (Word variations have been searched)

#26 (cystic fibrosis):ti,ab,kw AND (gastrointestinal):ti,ab,kw AND ("ultrasound scanning"):ti,ab,kw (Word variations have been searched)

#27 (cystic fibrosis):ti,ab,kw AND (gastrointestinal):ti,ab,kw AND ("computed tomography scan"):ti,ab,kw (Word variations have been searched)

#28 (cystic fibrosis):ti,ab,kw AND (gastrointestinal):ti,ab,kw AND ("X ray"):ti,ab,kw (Word variations have been searched)

#29 (cystic fibrosis):ti,ab,kw AND (gastrointestinal):ti,ab,kw AND (microbiota):ti,ab,kw (Word variations have been searched)

#30 (cystic fibrosis):ti,ab,kw AND (gastrointestinal):ti,ab,kw AND (terminal ileum motility):ti,ab,kw (Word variations have been searched)

#31 (cystic fibrosis):ti,ab,kw AND (gastrointestinal):ti,ab,kw AND (CFAbd-score):ti,ab,kw (Word variations have been searched)

#32 (cystic fibrosis):ti,ab,kw AND (gastrointestinal):ti,ab,kw AND (CFQoL):ti,ab,kw (Word variations have been searched)

#33 (cystic fibrosis):ti,ab,kw AND (gastrointestinal):ti,ab,kw AND (JenAbdomen-CF):ti,ab,kw (Word variations have been searched)

#34 (cystic fibrosis):ti,ab,kw AND (gastrointestinal):ti,ab,kw AND (PAC-SYM):ti,ab,kw (Word variations have been searched)

#35 (cystic fibrosis):ti,ab,kw AND (gastrointestinal):ti,ab,kw AND (PAC-QOL):ti,ab,kw (Word variations have been searched)

#36 (cystic fibrosis):ti,ab,kw AND (gastrointestinal):ti,ab,kw AND (PAGI-SYM):ti,ab,kw (Word variations have been searched)

#37 (cystic fibrosis):ti,ab,kw AND (gastrointestinal):ti,ab,kw AND (PAGI-QOL):ti,ab,kw (Word variations have been searched)

#38 (cystic fibrosis):ti,ab,kw AND (gastrointestinal):ti,ab,kw AND (GSRS):ti,ab,kw (Word variations have been searched)

39 (cystic fibrosis):ti,ab,kw AND (gastrointestinal):ti,ab,kw AND (PEI-Q):ti,ab,kw (Word variations have been searched)

#40 (cystic fibrosis):ti,ab,kw AND (gastrointestinal):ti,ab,kw AND (IBS-SS):ti,ab,kw (Word variations have been searched)

#41 (cystic fibrosis):ti,ab,kw AND (gastrointestinal):ti,ab,kw AND (abdominal symptom score):ti,ab,kw (Word variations have been searched)

### Clinical Trials Registries

#### ISRCTN
Searches:

- Cystic Fibrosis

#### Clinicaltrials.gov
Searches:

Cystic fibrosis, gastrointestinal
Cystic fibrosis, gastrointestinal symptoms
Cystic fibrosis, gastrointestinal dysfunction
Cystic fibrosis, gastrointestinal dysbiosis
Cystic fibrosis, meconium ileus
Cystic fibrosis, distal intestinal obstruction syndrome

### EU Clinical Trials Register

Searches:
Cystic fibrosis AND gastrointestinal
