## Supplementary material 3 for "Exploring the outcomes and endpoints used in gastrointestinal research in cystic fibrosis: a systematic review"

| GI subdomain | | |
|---|---|---|
| **GI growth and body composition** | **Number times used** | **Addition sub-domains mapped to** |
| Body mass index (BMI) | 37 | |
| Body mass index (BMI) z score | 35 | |
| Weight z score | 28 | |
| Height z score | 22 | |
| Weight | 11 | |
| Body mass index (BMI) percentile | 11 | |
| Fat free mass | 5 | |
| Height | 5 | |
| Weight percentile | 5 | |
| Fat mass | 4 | |
| Height percentile | 4 | |
| Length z score | 3 | |
| Length for age z score | 2 | |
| Weight for age z score | 2 | |
| Arm circumference percentile | 1 | |
| Arm circumference z score | 1 | |
| Arm muscle area | 1 | |
| Birth length z score catchup | 1 | |
| Birth weight z score catchup | 1 | |
| Body mass index (BMI) for age | 1 | |
| Body mass index (BMI) for age z score | 1 | |
| Fat area of the arm | 1 | |
| Fat mass index | 1 | |
| Fat mass percentile | 1 | |
| Fat mass z score | 1 | |
| Hand grip strength | 1 | |
| Head circumference percentile | 1 | |
| Lean tissue mass z score | 1 | |
| Length for age | 1 | |
| Peak height velocity | 1 | |
| Percentage body fat | 1 | |
| Percentage of lean mass | 1 | |
| Percentage of overweight or obesity | 1 | |
| Stature for age z score | 1 | |

| | |
|---|---|
| Tricipital skinfold measurement | 1 |
| Triciptal skin fold measurement z score | 1 |
| Weight for length percentile | 1 |
| Weight for length z score | 1 |

| **GI symptoms** | **Number times used** | **Addition sub-domains mapped to** |
|---|---|---|
| CFAbd-Score | 20 | Also maps to GI QoL |
| Patient Assessment of Constipation-Symptoms (PAC-SYM) | 14 | |
| Patient Assessment of Gastrointestinal Disorders-Symptom Severity Index (PAGI-SYM) | 7 | |
| GI symptom tracker | 4 | |
| IBS Severity Scoring System (IBS-SSS) | 3 | |
| Bristol stool chart | 2 | |
| GI symptom rating scale (GSRS) | 2 | |
| Number of stools | 2 | |
| Reflux symptom index (RSI) | 2 | |
| Reports of abdominal pain | 2 | |
| Brief pain inventory | 1 | |
| CFAbd-day2day | 1 | |
| Days of abdominal pain | 1 | |
| Fatigue assessment scale questionnaire | 1 | |
| Food diary myfood24 | 1 | |
| Frequency of abdominal symptoms | 1 | |
| Hull airways reflux questionnaire (HARQ) | 1 | |
| iCAN gastrointestinal tracker | 1 | |
| Incidence of constipation | 1 | |
| Medical symptoms reported in medical notes by pwCF or carers (reported as present or not present) | 1 | |
| Modified bristol stool chart (BSC) | 1 | |
| Number of stools per day | 1 | |
| Percentage prevalence of protocol described constipation | 1 | |
| PROMIS-GI | 1 | |
| Reporting of abdominal symptoms | 1 | |
| Reporting of selected gastrointestinal symptoms not part of a score | 1 | |
| Three constipation specific symptoms: report of incomplete bowel motions, straining to pass bowel motions and reported false alarm bowel motions | 1 | |
| Visual analogue score | 1 | |

| GI microbiota | Number times used | Addition sub-domains mapped to |
|---|---|---|
| Alpha diversity (Shannon index) | 11 | |
| Alpha diversity | 10 | |
| Beta diversity | 8 | |
| Bray curtis index | 7 | |
| Microbiota relative abudence | 4 | |
| Bacterial richness | 3 | |
| Faecal microbiota compostion | 3 | |
| Microbial diversity | 3 | |
| Phylum distribution | 3 | |
| Relative abudence of bacterial colonies | 3 | |
| Stool metabolomics | 3 | |
| Alpha diversity (simpsons index) | 2 | |
| Analysis of intestinal microbiota | 2 | |
| Dysbiosis score | 2 | |
| Fishers alpha index | 2 | |
| Microbiota richness | 2 | |
| Predtice functional analysis of bacterial present in the gut | 2 | |
| Abudence of specific amplicon sequence variants | 1 | |
| Analysis of compositions of microbiome | 1 | |
| Antibioticresistencegenes | 1 | |
| CAGS | 1 | |
| Camilleri or Ouyang motility indices and pressure curve | 1 | |
| Change in microbiome | 1 | |
| Changes in specific bacterial families (Bacteroidaceae and Porphyromonadacea) | 1 | |
| Detection of specific pathogens: staph aureus | 1 | |
| Diversity and composition of core and satellite taxa | 1 | |
| Genomospecies | 1 | |
| Glucose breath test | 1 | |
| Kegg orthologs and pathway analysis (relative abudence of gene orthologs using kegg protein database) | 1 | |
| Metagenomic analysis | 1 | |
| Microbial analysis | 1 | |
| Microbial pathways for aeorbic respiration | 1 | |
| Microbiome variability and abundance | 1 | |

| | |
|---|---|
| Pylotype richness | 1 |
| Relative abundance | 1 |
| Relative abundance of gammaproteobacteria | 1 |
| Relative microbiota age | 1 |
| Serum D-Lactate | 1 |
| Stool metagenomics | 1 |

| GI inflammation | Number times used | Addition sub-domains mapped to |
|---|---|---|
| Faecal calprotectin | 51 | |
| Faecal lactoferrin | 3 | |
| M2-Pyruvate Kinase | 3 | |
| Inflammatory markers including IL-1Beta, 6, 8, 10, 12p70 and TNF Alpha | 2 | |
| Dehydroepiandrosterone sulfate (DHEA-S) | 1 | |
| Faecal beta-defensin 2 | 1 | |
| Faecal myeloperoxidase | 1 | |
| Faecal neopterin | 1 | |
| Faecal osteoprotegerin | 1 | |
| Faecal S100A12 | 1 | |
| Histological evidence of intestinal mucosal inflammation | 1 | |
| Histological features of inflammation (rectal biopsy) | 1 | |
| Neutrophilic elastase | 1 | |
| Serum Intestinal-Fatty Acid Binding Protein (I-FABP) | 1 | |

| Gi motility | Number times used | Addition sub-domains mapped to |
|---|---|---|
| Small bowel water content (SBWC) | 7 | |
| Orocaecal transit time (MRI) | 6 | |
| Delta small bowel water content (SBWC) | 4 | |
| Gastric half empty time (MRI) | 2 | |
| Number of contractions of colon (wireless motility capsule) | 2 | |
| Small bowel transit time (endoscopic capsule) | 2 | |
| Total and segmental colonic transit time | 2 | |
| Ascending colon longitudinal relaxation time | 1 | |
| Delayed gastric emptying (gastric emptying scintigraphy) | 1 | |
| Endoluminal motion (wireless motility capsule) | 1 | |
| Gastric empty (gamma camera) | 1 | |

| | |
|---|---|
| Gastric emptying rate (using 13C-octanoate breath test) | 1 |
| Gastric transit time | 1 |
| Gastrointestinal transit time (capsule) | 1 |
| Ileocoloic/ gastrointestinal transit time | 1 |
| Images containing turbid luminal contents (endoscopic capsules) | 1 |
| Number of luminal closures (measure of contractility, endoscopic capsule) | 1 |
| Retention of turbid material (wireless motility capsule) | 1 |
| Small bowel motility | 1 |
| Small bowel transit time | 1 |
| Small bowel transit time (gamma camera) | 1 |
| Terminal ileum motility score | 1 |
| Transit time | 1 |

| **GI Quality of life** | **Number times used** | **Addition sub-domains mapped to** |
|---|---|---|
| Cystic Fibrosis Questionnaire - Revised (CFQR) | 16 | |
| PedsQL Gastrointestinal Symptoms Scales 3.0 | 6 | Also GI symptoms |
| Patient Assessment of Constipation Quality of Life questionnaire (PAC-QOL) | 5 | |
| Cystic Fibrosis specific Pediatric Quality of Life Inventory Gastrointestinal Symptoms Module (CF-PedsQL-GI) | 3 | Also GI symptoms |
| PedsQL Generic Core Scale 4.0 | 3 | |
| Gastrointestinal quality of life index (GIQLI) | 1 | |
| Intestinal wellbeing evaluated by 7 item questionnaire (based on 7 variables diarrhoea, upper and lower abdominal pain, abdominal swelling, flatulence, constipation, hyperoxia | 1 | |

| **Pancreatic exocrine insufficiency** | **Number times used** | **Addition sub-domains mapped to** |
|---|---|---|
| Faecal elastase | 25 | |
| Faecal steatocrit | 3 | |
| Faecal chymotrypsin | 1 | |
| Faecal fat profiling | 1 | |
| Lipase substitution dose | 1 | |
| Pancreastic liapse actibity via 13C triglyceride breath test | 1 | Also GI absorption |
| Pancreatic secretion rate (i.e. secretions within 1-5 minutes of secretin stimulation) | 1 | |
| Secretin stimulated peak values measured using sonography | 1 | |
| Secretin-stimulated intestinal fluid volume change | 1 | |
| Secretin-stimulated pancreatic fluid volume secretion (using s-MRI) | 1 | |

| | | |
|---|---|---|
| Serum immunoreactive trypsinogen | 1 | |
| Short endoscopic secretin test to measure: duodenal bicarbonate, lipase, amylase, elastase, chymotrypsin and aspirated volume of fluid | 1 | |
| Subjective assessment of of stool fat and consistency | 1 | |
| Total secreted volume from pancreas (change in small bowel volume pre and post secretin stimulation) | 1 | |
| **Gastrointestinal absorption, digestion, and emulsification** | **Number times used** | **Addition sub-domains mapped to** |
| Coeffiecient of fat absorption | 9 | |
| Coefficient of nitrogen absorption | 3 | |
| Essential fatty acids | 3 | |
| Short chain fatty acid profiling | 3 | |
| Gut permeability (urinary lactulose/mannitol) | 2 | |
| Serum levels of fat-soluble vitamins | 2 | |
| Acid steatocrit | 1 | Also PEI |
| Albumin | 1 | |
| Colon chyme Haralick contrast (MRI) | 1 | Also intraluminal environment |
| Fatty acid profile | 1 | |
| Linoleic acid | 1 | |
| Malabsorption blood test | 1 | Also PEI |
| Prealbumin | 1 | |
| Small bowel chyme texture | 1 | Also intraluminal environment |
| Stool fat loss | 1 | |
| Total protein values | 1 | |
| Vitamin D level | 1 | |
| **Oesophageal and GI intraluminal environment** | **Number times used** | **Addition sub-domains mapped to** |
| pH measurement (wireless motility capsule) | 2 | |
| pH profile area under the curve | 2 | |
| Bicarbonate concentration (wireless motility capsule) | 1 | Also PEI |
| Bowel compressibility and bowel content characteristics (US) | 1 | |
| Colon mucosal appearance | 1 | |
| DeMeester Score (oesophageal pH monitoring and multichannel intraluminal impedance) | 1 | |

| | |
|---|---|
| Expression of colonic acid-base transporters | 1 |
| Gastrointestinal pH | 1 |
| Intestinal current measurement | 1 |
| Mean duration of acid events (oesophageal pH monitoring and multichannel intraluminal impedance) | 1 |
| Mean pH between 8 and 24 minutes from gastric emptying | 1 |
| Mean pH values | 1 |
| Mean pH values 23 minutes from gastric emptying | 1 |
| Mean time required to reach and sustain pH of 5.5 | 1 |
| Number of acid events (oesophageal pH monitoring and multichannel intraluminal impedance) | 1 |
| Number of events lasting >5 minutes (oesophageal pH monitoring and multichannel intraluminal impedance) | 1 |
| Reflux index | 1 |
| Slope comparison of changes in pH profile | 1 |
| Time interval required to reach and maintain pH of 5.5 and 6.0 | 1 |
| Time taken to sustain a mean pH >5.5 | 1 |

| **GI structure** | **Number times used** | **Addition sub-domains mapped to** |
|---|---|---|
| Colonic volume (MRI) | 4 | Also GI motility |
| 2D shear wave elastrography | 3 | Also pancreatic structure |
| Multiple ultrasound findings including: bowel wall thickness, mesenteric lymph nodes, intussusception, appendiceal thickening, free fluid with the peritoneal cavity, pancreatic cystosis and pancreatic lipomatosis | 2 | |
| Area of the descending duodenum | 1 | |
| Bowel wall thickness, bowel wall pattern and vascularisation within the bowel wall (US) | 1 | |
| Composite frequency (score) of symptoms per abdominal ultrasound abnormality | 1 | Also GI symptoms |
| Diameter of the descending duodenum | 1 | |
| Ileal perimeter (MRI) | 1 | |
| Luminal volumes of the ileum and colon (MRI) | 1 | |
| Measurement of ultrasound abnormalities | 1 | Also pancreatic structure |
| Mesenteric adipose tissue alteration, enlarged mesenteric nodes (US) | 1 | |
| Number and severity of intestinal lesions using maiden classification score, captured via video capsure endoscopy | 1 | |
| Small bowel loop diameter (using ultrasound) | 1 | |
| Terminal ileum diameter (MRI) | 1 | |

| Complications of CF | Number times used | Addition sub-domains mapped to |
|---|---|---|
| Incidence of meconium ileus | 2 | |
| Prevalence of meconium ileus | 2 | |
| Bowel motility, luminal narrowing or stenosis or intussusception (US) | 1 | Also GI motility |
| Faecal impaction | 1 | |
| Incidence of DIOS | 1 | |
| Incidence of gastrointestinal cancer | 1 | |
| Incidience of pancreatic insufficiency | 1 | Also PEI |
| Intestinal manifestations in CF | 1 | |
| Meconium ileus phenotype (MIP) score | 1 | |
| Pancreatic insufficiency phenotype score | 1 | Also PEI |
| Pancreatic manifestations of CF | 1 | |
| Rate of distal intestinal obstruction syndrome (DIOS) | 1 | |
| Rate of gastroesophageal reflux disease | 1 | |
| Rate of pancreatic insuffiency | 1 | Also PEI |
| Rate of pancreatic exocrine insufficiency | 1 | Also PEI |

| Pancreatic structure | Number times used | Addition sub-domains mapped to |
|---|---|---|
| Pancreatic apparent diffusion coefficient | 2 | |
| Visual assessment score of pancreatic hyperechogenicity | 2 | |
| Appearance of the general structure of the pancreas using ultrasound | 1 | |
| Diameter of anatomical parts of the pancreas (ultrasound) | 1 | |
| Diameter of the wirsung duct | 1 | |
| Pancreatic fat-signal fractions and fat/water ratios | 1 | |
| Pancreatic gland volume | 1 | |
| Pancreatic size | 1 | |
| Pancrenchymal and ductal characteristics on ultrasound | 1 | |
| Visual assessment score between pancreatic echogenicity compared to the liver | 1 | |

| CF medication use | Number times used | Addition sub-domains mapped to |
|---|---|---|
| Dose and frequency of PERT use | 1 | |
| Lipase dose | 1 | |
| Number capsules of PERT | 1 | |

| | | |
|---|---|---|
| Rate of fat soluble vitamin use | 1 | |
| Rate of Pancreatic Enzyme Replacement Therapy (PERT) use | 1 | |
| Reported use of acid suppression medication | 1 | |
| **Diet and dietary change** | **Number times used** | **Addition sub-domains mapped to** |
| Australian child and adolescent eating survey | 3 | |
